# Assessments of Effectiveness of Technologies Utilizations in VIHSCM Among Selected Health Facilities in Tanzania Mainland

**DOI:** 10.1101/2023.10.31.23297838

**Authors:** Henry A. Mollel, Lawrencia D. Mushi, Richard V. Nkwera

## Abstract

**Introduction:** Tanzania has adopted various technologies for Vaccine and Immunization Health Supply Chain Management to improve the availability, access, and utilization of immunization programs. However, questions remain regarding the effectiveness of the technologies in Vaccine and Immunization Health Supply Chain Management. This study assesses the effectiveness of technology on vaccine and immunization supply chain management in selected health facilities in Tanzania.

**Methods:** This study adopted an exploratory descriptive cross-sectional design. The study collected data using structured questionnaires from health facilities that adopted VIHSCM technologies in Arusha, Mwanza, and Mbeya regions, Tanzania. Data were analyzed using descriptive statistics and cross-tabulations with the aid of the Statistical Package of Social Sciences 23^rd^ Edition (SPSS).

**Results:** The study findings showed that 56.7% of the surveyed Health facilities had either TiMR, DHIS2, or GOTHOMIS technology used for vaccine and immunization supply chain management. The study shows that 51.4% of respondents at health facilities agreed that the adopted technologies were very effective, 45.9% were moderately effective and 2.7% said the technologies were less effective. The results asserted that 18.9 % of the adopted technologies were effective in the Management of immunization services, 37.8 % asserted that technology utilizations have Increased access to the vaccine, 2.7% said that technology utilizations assisted in monitoring vaccine temperature, 24.3% said that technologies utilizations assisted in vaccines at their respective health facilities, 5.4 % said that technologies utilizations assisted in the Maintaining the quality of Vaccines.

**Conclusion:** This study concluded that technology plays a substantial role in improving the availability and quality of vaccines and immunization services in health facilities. Also, the study suggested that increase the use of technologies to capitalize the VIHSCM effectiveness.

## 1. Introduction

Immunization Supply Chain and Logistics is the key area for the Immunization Agenda 2030 (IA2030) (1). IA2030 focuses on ensuring that immunization programs are an integral part of primary health care to achieve universal health coverage (PHC), Universal Health Care (UHC), and the health Sustainable Development Goal (SDG) for the decade 2021-2030 (2). In Africa, immunization programs have played a key role in successfully strengthening African immunization systems to sustainably and equitably increase immunization coverage rates and bolster health system performance at the national and sub-national levels (3). According to the Expanded Programme on Immunization (EPI) by providing universal access to immunization, the region has achieved a dramatic reduction in infant and under-five mortality in Africa in the last few decades(4). The success of EPI rests on the national vaccine supply chain systems which are constantly under increasing pressure to ensure that high-quality vaccines are always available in the right quantity and form at the right time, in the right place, and stored and distributed under the right conditions (1, 4). Over a decade, routine immunization coverage in the United Republic of Tanzania has increased considerably, from 86% to 98% coverage of the third dose of the diphtheria-tetanus pertussis (DTP) vaccine, and from 88% to 99% coverage of the first dose of the measles-rubella vaccine (5). To maintain these Achievements and continue scaling up newer vaccines, Tanzania’s immunization program will need sufficient and effective technologies to ensure reliable, strong and efficient health supply chain system to deliver immunization services to achieve intended health outcomes (6, 7). Targeting to improve the performance of vaccines and immunization services in the country Tanzania’s Ministry of Health (MoH) through its Expanded Program for Immunization (EPI) program adopted vaccines and immunizations health supply chains management (VIHSCM) technologies and innovation such as Vaccine Information Management System (VIMS), Warehouse Management Information systems (Part of the VIMS Module) and Tanzania Immunization Registry (TImR) (8, 9). The adopted technologies have an essential role in strengthening the management of vaccine and logistic information at the health facility, solving challenges such as inadequate data management and use capacity at all levels of the health system, poor data visibility into supplies at the facility level to district level data and stock challenges as a result of insufficient supply chains and logistics management.

Despite the initiatives by the Government with facilitations from development partners there exist scant evidence on performance of VIHSCM technologies in Tanzania. Previous observational studies such as (10) assessed vaccine management practices among vaccinators at health facilities in the Morogoro region, Tanzania, (11) assessed the effect of electronic immunization registry practices on the availability of immunization commodities while (12) assessed trends in immunization completion and disparities in the context of health reforms in Tanzania, (13) evaluated the impact of upgrading supply chain management systems to improve availability of Medicines in Tanzania; (14) compared the actual storage and distribution costs of vaccines and related supplies between MSD to EPI and Immunization coverage country reports (1, 15). These studies affirmed that prevalence of inequities and disparities in immunization coverage across districts and health facilities level due to lack of electricity, poor network connectivity, and change of users with lower digital literacy. Furthermore, previous studies show that most of these adopted technologies and innovation have only benefited part of the population. Therefore, this study aimed to assess the effectiveness of existing technologies in VIHSCM systems across health facilities within Tanzania’s health system along with describing aspects in which adopted technologies has improved access and utilizations of vaccines and immunization for children in Tanzania.

## 2. Design and Method

### 2.1 Study Setting

A facility based exploratory descriptive cross-sectional design with mixed methods approach was employed to understand the effectiveness of utilizations of technologies for VIHSCM across health facilities in three purposively selected regions of mainland Tanzania namely Arusha, Mwanza, and Mbeya. This research design allowed both qualitative and quantitative data to be collected simultaneously, analyzed separately, and later combined for interpretation “in order to provide a comprehensive analysis of the effectiveness of utilizations of technologies for VIHSCM across health facilities [15].

### 2.2 Data Collection Instruments and sampling procedures

Data collection was conducted in three purposively selected regions of mainland Tanzania namely Arusha, Mwanza and Mbeya. In each of the selected region, two districts were purposively selected. The criteria for selection of both regions and districts was urban-rural divide and the rate of vaccines and immunization uptake rate for the period of 20^th^ January 2022 to 19^th^ January 2023. The selection was based on capturing contextual variations that can influence the uptake, scale up and integration of adopted technologies and innovations into existing systems and policies. Key informants were selected at all levels based on their involvement in the supply chain management.

In the selected regions and councils, two and three relevant officials were selected purposively on the basis of involvement in the implementation of vaccines and immunization activities. These included the Regional Medical Officer, Regional Immunization and Vaccine Officer, District Medical Officer, District Immunization and Vaccine Officer, and District HMIS (MTUHA) Focal Person. In each of the selected health facility, three staff were purposively selected based their involvement in vaccine and immunization management. These were-health facility in-charge, the in-charge of Reproductive and Child Health (RCH), and RCH vaccine coordinator. It is important to notice that in some health facility the in-charge of RCH served also as the vaccine coordinator.

### 2.3 Data collection

Primary data were collected using semi-structured interview guide, questionnaire and observations checklist developed by reviewing early studies. Secondary data were collected through documentary review. Interviews was conducted to 41 key informants at national, regional and council levels. Questionnaire was administered to 37 respondents at health facility level. Interviewed guide was also used to collect data from one group interview in the Faith Based Owned health facility and one Focus Group Discussion in the private health facility.

### 2.4 Data Processing and Analysis

Data collected during the study was cleaned, processed and analyzed using IBM Statistical Package for Social Studies (SPSS). Because of the small number of records, the analysis only focus on descriptive statistics (frequencies, percentages, means, and standard deviation), as well as chi-square tests were performed for all demographic and health facilities characteristics as well as for study variables including health workers understanding on presence and utilizations of technologies for VISHSCM.

### 2.5 Ethical considerations

Ethical clearance approval (NIMR/HQ/R.8a/Vol.IX/3878 of 20th January 2022) was obtained from the National Institute for Medical Research under the Tanzania Ministry of health. The research was also approved by Mzumbe University. Permission to conduct research in the relevant institutions were obtained from the Ministry of Health (MOH) and President’s Office Regional Administration and Local Government (PORALG). Respondents were orally informed of the purpose of the research and provided a consent form to sign after voluntarily agreeing to participate. The participants’ confidentiality was maintained throughout the study, including not exposing their personal identities or names of facilities at which they were working.

## 3. Results

### 3.1 Health Facility and Respondents Characteristics

From 37 health facilities enrolled, 37 gave complete response making response rate 100 percent. The study assessed health facilities in four regions of Arusha, Morogoro, Mbeya, and Mwanza regions. A total of 4 regions 8 districts and 37 health facilities participated in this study. Out of the studied health facilities 22 (59.5%) were District hospitals, 4 (10.8%) were Health centers and 11 (29.7 %) were dispensaries. The study shows that of 11 (29.7 %) respondents were male while 26 (70.3%) of the respondent were female. The age distribution of the respondent across health facilities showed that the average age was 45 years old. Out of 37 respondents 12(32.4%) were at the age group between 25 and 34 years; 12(32.4%) were at the age group between 35 and 44 years, 10 (27%) were at the age group between 45 and 54 years; 3 (8.1%) were at the age group between 55-64 years. The academic qualifications of respondents were as follows. 2 (5.4%) of respondents had an advanced diploma, one in clinical medicine working at dispensary and the other in nursing working at the district hospital. About 8 (21.6%) respondents in the health facility had certificates and qualifications in clinical medicines and nursing, 4 working at a dispensary, 3 working at a District hospital and one working at the health Centre. About 15 (40.5%) respondents in the health facility had diploma qualifications in clinical medicines and nursing, 5 working at dispensary level, 10 working at District hospital. About 9 (21.6%) respondents in the health facility had medical doctor qualifications, 7 working at District hospital and 2 working at health center. More results on Demographic and health facilities characteristics are found in Table 1.

**Table 1:** Demographic and Facilities characteristics across Health Facility Level.

| Variables | Status | Health Facility Level (%) |  |  | Total<br>(n=37) |
| --- | --- | --- | --- | --- | --- |
|  |  | District<br>Hospital | Health<br>Centre | Dispensary |  |
| Sex of<br>respondent | Male | 7 (31.8) | 2 (50) | 2 (18.2) | 11(29.7) |
|  | Female | 15 (68.2) | 2 (50) | 9 (81.8) | 26(70.3) |
| Age of<br>respondent | 25-34 | 7 (31.8) | 0 (0%) | 5(45.5) | 12(32.4) |
|  | 35-44 | 6 (27.3) | 2 (50) | 4 (36.4) | 12(32.4) |
|  | 45-54 | 8 (36.4) | 1 (25) | 1 (9.1) | 10(27) |
|  | 55-64 | 1 (4.5) | 1 (25) | 1 (9.1) | 3(8.1) |
| Job title | Facility<br>incharge | 0 (0%) | 0 (0%) | 1 (9.1) | 1(2.7) |
|  | Facility RCH<br>incharge | 3 (13.) | 1 (25) | 5 (45.5) | 9(24.3) |
|  | Facility RCH<br>nurse | 0 (0%) | 0 (0%) | 1 (9.1) | 1(2.7) |
|  | Facility RCH<br>vaccine<br>coordinator | 7 (31.8) | 0 (0%) | 4 (36.4) | 11(29.7) |
|  | Medical facility<br>incharge | 0 (0%) | 2 (50) | 0 (0%) | 2(5.4) |
|  | Medical officer<br>incharge | 6 (27.3) | 1 (25) | 0 (0%) | 7(18.9) |
|  | MTUHA | 6 (27.3) | 0 (0%) | 0 (0%) | 6(16.2) |
|  | Advanced<br>diploma in<br>nursing | 1 (4.5) | 0 (0%) | 0 (0%) | 1 (2.7) |
|  | Assistant<br>medical officer | 0 (0%) | 0 (0%) | 1 (9.1) | 1 (2.7) |
|  | Certificate in<br>clinical<br>medicines | 0 (0%) | 0 (0%) | 1 (9.1) | 1 (2.7) |
|  | Certificate in<br>nursing | 3 13.6) | 1 (25) | 3 (27) | 7 (18.9) |
|  | Clinical officer | 1 (4.5) | 0 (0%) | 0 (0%) | 1 (2.7) |
|  | Diploma in<br>clinical<br>medicines | 2 (9.1) | 0 (0%) | 2 (18.2) | 4 (10.8) |
|  | Diploma in<br>nursing | 8 (36.4) | 0 (0%) | 3 (27.3) | 11 (29.7) |
|  | Medical doctor | 7 (31.8) | 2 (50) | 0 (0%) | 9 (24.3) |
|  | Surgeon | 0 (0%) | 1 (25) | 0 (0%) | 1 (2.7) |
| Duration in<br>current post<br>(Groups) | <1 Years | 2 (9.1) | 0 (0%) | 2 (18.2) | 4 (10.8) |
|  | 1-5 Years | 11 (50) | 0 (0%) | 6 (54.5) | 17 (45.9) |
|  | 6-10 Years | 7 (31.8) | 2 (50) | 1 (9.1) | 10 (27) |
|  | >11 Years | 2 (9.1) | 2 (50) | 2 (18.2) | 6 (16.2) |

### 3.2 Status of Technology Utilizations for VIHSCM across health facilities

The rapid advancement in utilizations of technology in vaccines and immunizations supply chains management provides an important opportunity to ensure that each health facility receives its vaccines and inputs at the right time, in the right amount, in the right conditions and temperatures(16). The results in Table 2 shows that all of 37 visited health facilities six in visited district councils reported presence of refrigerators. Refrigerators are critical components in the management of the vaccines quality(17). The results showed that software and forms are used to report children utilizations of vaccines and immunizations in health facilities. The results show that 4 (10.8%) health facilities used Electronic software where as 3 were district hospital, 1 Health Centre. The results show that 18 (48.6%) used forms to report vaccines utilizations where as 9 District hospitals, 1 health center and 8 dispensaries. The results show that 15 (40.5%) health facilities used both Electronic software and forms where as 10 were district hospital, 2 Health Centre and 3 dispensaries.

**Table 2:** Technology utilized for vaccines and immunizations management across health facilities.

|  | Technologies | District Hospital | Health Centre | Dispensary | Total |
| --- | --- | --- | --- | --- | --- |
| **Technology used for vaccines and immunization storage (B5)** | **Refrigerator** | 22(59.5%) | 4(10.8%) | 11(29.7%) | 37 (100%) |
| **What are you using to report the used vaccines?(B4)** | **Software** | 3 (13.6%) | 1 (25%) | 0 | 4 (10.8%) |
|  | **Form** | 9 (40.9%) | 1 (25%) | 8(72.7%) | 18 (48.6%) |
|  | **Both software and form** | 10 (45.5%) | 2 (50%) | 3 (27.3%) | 15 (40.5%) |
|  | **Total** | 22 (100%) | 4(100%) | 11(100%) | 37(100%) |

### 3.3 Tools for Vaccines and Immunization Management

The results showed that in order to administer vaccines for children immunizations the health facilities are using Electronic software and forms to administering vaccines and immunizations in health facilities. The results in Table 3 show that of 5 (13.5%) health facilities used Electronic software where as 2 were district hospital, 1 Health Centre and 2 dispensaries. The results show that 18 (48.6%) used forms to administer vaccines where as 10 District hospitals, 2 health center and dispensaries. The results show that 14 (37.8%) health facilities used Electronic software and forms where as 10 were district hospital, 1 Health Centre and 3 dispensaries.

**Table 3:** Technology for Vaccines and Immunization Management across health facility.

|  | **Technology** | **District Hospital** | **Health Centre** | **Dispensary** | **Total** |
| --- | --- | --- | --- | --- | --- |
| **Tool used to administer vaccine (B10)** | **Electronic software** | 2(9.1%) | 1(25%) | 2(18.2) | 5(13.5) |
|  | **Form** | 10(45.5%) | 2(50%) | 6(54.4%) | 18(48.6%) |
|  | **Both electronic software and form** | 10(45.5%) | 1(25%) | 3(27.3%) | 14(37.8%) |
|  |  | 22(100) | 4(100) | 11(100) | 37(100) |
| **VIHSCM Tool for Supplies & Logistics Mgmt (B10)** | **Integrated Logistics Systems (ILS)** | 0 | 1 (25%) | 0 | 1 (2.7%) |
|  | **Tanzania Immunization Registry (TImR)** | 10(45.5%) | 0 | 3(27.3%) | 13 (35.1%) |
|  | **DHIS 2** | 1(4.5%) | 0 | 2(18.2%) | 3(8.1%) |
|  | **GoTHOMIS** | 4(18.2) | 0 | 1(9.1%) | 5(13.5%) |
|  | **No software** | 7(7) | 3(75%) | 5(45.5%) | 15(40.5%) |
|  |  | 22(100) | 4 (100) | 11(100) | 37(100) |

The results from the study showed that for managing supplies and logistics of vaccines and immunizations services Integrated Logistic System (ILS) was only utilized at 1 (2.5%) Health center, the Tanzania Immunization Registry (TImR) was utilized at 13 (35.1%) health facilities that is 10 (45.5%) District Hospital and 3 (27.3%) Dispensaries. Furthermore, the results revealed that the DHIS2 was utilized at 3 (8.1%) of surveyed health facilities that is 1 (4.5%) District hospitals and 2(18.2%) Dispensaries, GoTHOMIS was utilized at 5 (13.5%) of health facilities that is 4 (18.2%) District hospitals and 1 (9.1%) being a Dispensary. The study revealed that 15 (40.5%) of surveyed health facilities had no technology/ software tools to facilitate supplies and logistics management of VIHSCM that is 7 District hospital, 3 and 5 Dispensaries.

### 3.4 Relevancy and Effectiveness of Technology utilizations in VIHSC Management

The results in Table 4 shows that from the 37 surveyed Health facility 7 (18.9%) of the respondent asserted that technology has assisted to increase access and utilizations of vaccines and immunizations across 4 District hospitals and 3 Dispensary. About 14 (37.8%) respondents at health facilities asserted that technology utilization has increased access of vaccines and immunizations services for children’s across 8 district hospitals, 2 Health centers, and 4 dispensary ; About 9 (24.3%) of respondents at health facilities asserted that technology utilization has increased availability of vaccines and immunizations services for children’s across 6 district hospitals, 2 health center and 1 dispensary ; About 1 (2%) of health facilities asserted that technology utilization has increased monitoring of vaccines and immunizations ordering. 2 (5.4%) of health facilities which were District hospitals asserted that technology has assisted in maintaining of quality of vaccines, 2 (5.4%) of health facilities which were Dispensaries’ asserted that technology has assisted in monitoring of vaccines temperature, 2 (5.4%) health facilities which were District hospitals asserted that technology utilization facilitated in providing direct communication between health facility and DIVO.

**Table 4:**
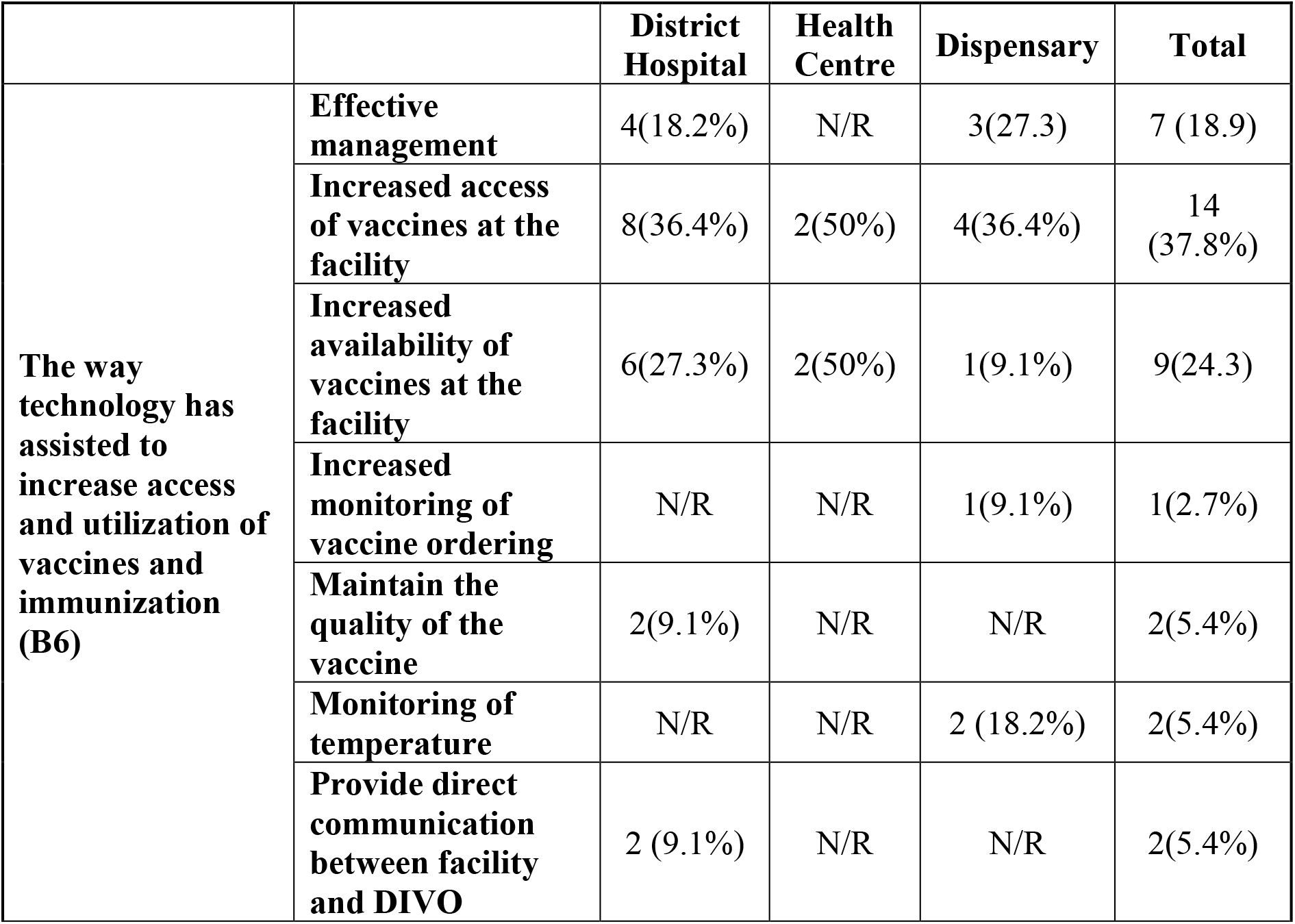
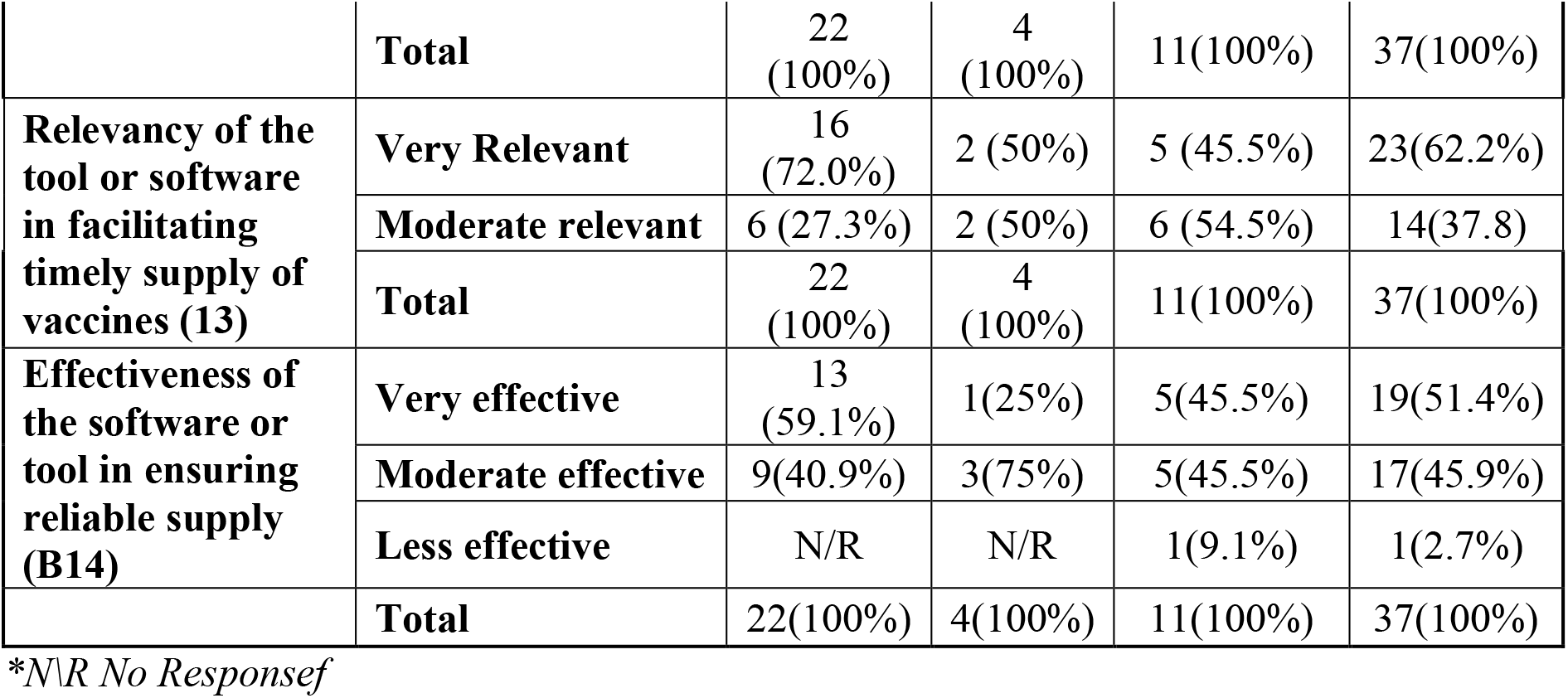
Effectiveness of Technology utilizations in VIHSC Management across health facilities.

|  |  | **District Hospital** | **Health Centre** | **Dispensary** | **Total** |
| --- | --- | --- | --- | --- | --- |
| **The way technology has assisted to increase access and utilization of vaccines and immunization (B6)** | **Effective management** | 4(18.2%) | N/R | 3(27.3) | 7 (18.9) |
|  | **Increased access of vaccines at the facility** | 8(36.4%) | 2(50%) | 4(36.4%) | 14 (37.8%) |
|  | **Increased availability of vaccines at the facility** | 6(27.3%) | 2(50%) | 1(9.1%) | 9(24.3) |
|  | **Increased monitoring of vaccine ordering** | N/R | N/R | 1(9.1%) | 1(2.7%) |
|  | **Maintain the quality of the vaccine** | 2(9.1%) | N/R | N/R | 2(5.4%) |
|  | **Monitoring of temperature** | N/R | N/R | 2 (18.2%) | 2(5.4%) |
|  | **Provide direct communication between facility and DIVO** | 2 (9.1%) | N/R | N/R | 2(5.4%) |
|  | **Total** | 22<br>(100%) | 4<br>(100%) | 11(100%) | 37(100%) |
| **Relevancy of the tool or software in facilitating timely supply of vaccines (13)** | **Very Relevant** | 16<br>(72.0%) | 2 (50%) | 5 (45.5%) | 23(62.2%) |
|  | **Moderate relevant** | 6 (27.3%) | 2 (50%) | 6 (54.5%) | 14(37.8) |
|  | **Total** | 22<br>(100%) | 4<br>(100%) | 11(100%) | 37(100%) |
| **Effectiveness of the software or tool in ensuring reliable supply (B14)** | **Very effective** | 13<br>(59.1%) | 1(25%) | 5(45.5%) | 19(51.4%) |
|  | **Moderate effective** | 9(40.9%) | 3(75%) | 5(45.5%) | 17(45.9%) |
|  | **Less effective** | N/R | N/R | 1(9.1%) | 1(2.7%) |
|  | **Total** | 22(100%) | 4(100%) | 11(100%) | 37(100%) |

\*N\ R No Response

More over in terms of the relevance of technology in facilitating timely Supply of vaccines for children immunizations across health facilities in the country 23 (62.2) of health facilities that is 16 district hospitals, 2 Health centers and 5 Dispensary found technology to be very relevant, 14 (37.8) of health facilities that is 6 district hospitals, 2 Health center, and 6 Dispensary found technology to be moderate relevant

Moreover, in terms of the effectiveness technology in ensuring a reliable supply of vaccines for children immunizations across health facilities in the country 19 (51.4) of health facilities that is 13 district hospitals, 1 Health centers and 5 Dispensaries found technology to be very effective, 17 (45.9) of health facilities that is 9 district hospitals, 3 Health center, and 5 Dispensary found technology to be moderate effective. 1 (2.7) of health facilities that is 1 Dispensary found technology to be less effective in ensuring a reliable supply of vaccines for children immunizations across health facilities in the country.

### 3.5 Tendencies of Vaccines stock out and ordering across health facilities levels

The respondents in the study were asked on their view if their facilities have experience stocks out and how long does it take do reordering and delivering of vaccines at health facilities. The results in Table 5 show that 18 (48.6) of health facilities that is 9 District hospital, 2 Health center and 7 Dispensary respectively of health facilities in the study experienced stock out once in year and 19 (51.4%) of health facilities in the study that is 13 District hospital, 2 Health center and 4 Dispensary respectively never experienced stock out. The results shows that the adopted technology were effective in reducing stock out rates and ensuring time delivery of Immunizations services, same results were reported by (13).

**Table 5:** Tendencies of Vaccines stock out and ordering across health facilities levels.

|  |  | District Hospital | Health Centre | Dispensary | Total |
| --- | --- | --- | --- | --- | --- |
| How often do you get out of stock for the specified vaccines? | Once a year | 9(40.9) | 2 (50) | 7(63.6) | 18(48.6) |
|  | Never experienced stock-out | 13(59.1) | 2 (50) | 4(36.4) | 19(51.4) |
| How long does it take from ordering to delivery? | Takes only few days | 22 (100) | 4(100) | 11(100) | 37(100) |

## 4. Discussion

This study highlighted the utilization of technology in children vaccine and immunizations supply chain management using both interview-based and actual-practice observational studies. The findings in **Error! Reference source not found**.2 confirms that health workers are using technologies for storing vaccines and reporting utilizations of Vaccines in the health facilities. The Technologies have been useful in facilitating reliable availability of vaccines. The results show that most of the Health facilities are transitioning from paper work to paperless in ordering, administering and reporting. As indicated from the results most have not experienced stock-out for a while and only reported to experience stock-out once a year. The quality of vaccines has been managed thought effective use of refrigerator and Remote Temperature Monitoring technology. The technology was reported by respondents to be useful in ensuring adherence of managing the temperature at required range and maintain its condition for safety use (cold chain system). The use of technology has also enabled feedback of vaccinated individuals and their locations which has been useful in developing strategies of how to reach more population in case of under vaccination reports.

The utilizations of Vaccines and Immunization of Health Supply Chain Management technologies as indicated in **Error! Reference source not found**.3, the column of ILS (integrated logistic system) is reported zero because the target respondents (i.e. RCH in-charge, dispensing nurse, MTUHA focal person) are not using that system in their routine services and the vaccines are separated from pharmaceuticals. TImR is reported to some facilities because this system is not scaled up countrywide. It is only rolled in 3736 health facilities in 15 (57.7%) regions. In the studied regions, TImR was only rolled out in Arusha and Mwanza. It was not yet rolled out in Mbeya. It was also learned that, in Meru and Longido District Councils there were good number of broken tablets that led to stoppage use of TIMR system. As further indicated in Table 3, TIMR in used in both public and private health facilities. GoTHOMIS is only used at District Council hospital levels. It not yet rolled out at levels of Health Centre and Dispensary.

Yet data on vaccines and immunization are still filled out manually and electronic, indicating that the technology is still not fully in utilizations as indicated in Table 2. Although all studied health facilities reported availability of functional refrigerators, Remote Temperature Monitoring (RTM) technology is only used at national, regional and council levels. In view of most respondents, RTM is a most useful technology that need to be scaled up to health facility levels.

## 5 Conclusion

Overall this study aimed to assess the utilization of technology in children vaccine and immunizations supply chain management using both interview-based and actual-practice observational studies. The study concluded that VIHSCH technologies are recognised being effective to improve availability and quality of vaccines and immunization services in health facilities. The Technologies has reduced stock out incidence and improved delivery of Immunizations commodities. There was high recognition among participants to on the utilizations of Vaccine Information Management System (VIMS), Warehouse Management Information systems, Tanzania Immunization Registry (TImR) and Storage Technologies-in facilitating quality and access of vaccines and immunization services. Despite the recognition-there were concerns on the adoption, design and institutional factors that support the effectiveness of technology utilizations for Vaccines and immunizations of health supply chain management among health facilities in regions across Tanzania.

## Data Availability

The data sets used and/or analyzed during the current study are available from the corresponding author on reasonable request.

## 6 Research Limitations

Data collections was conducted in three regions and six councils and therefore our findings, conclusion and recommendations reflect the settings and technologies applied in those councils.. Using a small sample size might make it difficult to draw conclusions and generalize. However, since the context in which Vaccination and Immunization of VIHSCM processes do not vary significantly and guided by similar operational framework, the findings can be replicated to other contexts. Despite these limitations, the study’s direct observation study design can show an actual practice. vaccine and cold chain management practices. Future research needs to be conducted with larger sample of health facilities and health workers in the country so as to be able to realise and generalise the effectiveness of technologies utilisation technologies for VIHSCM.

### List of Abbreviation

RCH: Reproductive and Child Health
RTM: Remote Temperature Monitoring
GoTHoMIS: Government of Tanzania Health Operation Management Information System
TImR: Electronic Immunization Registry
VIHSCM: Vaccines, Immunization and Health Supply Chain Management
VIMS: Vaccine Information Management System
UNICEF: United Nations International Children’s Emergency Fund

## Declarations

### Consent for publication

Not Applicable

### Authors’ Contributions

Henry A. Mollel, Lawrencia D. Mushi, and Richard V. Nkwera have contributed to data collection and data analysis. All authors commented on previous versions of the manuscript, read and approved the final manuscript.

## Acknowledgements

This research was funded by UR, EAC Regional Center of Excellence for Vaccines, Immunisation, and Health Supply Chain Management (UR EAC RCE-VIHSCM) through the Research Grant Schemes. The authors thank the top leadership of Mzumbe University for their technical support and in facilitating collaboration with the University of Rwanda and implementation of this study. We are also thankful to Noel Otieno, Irene Moshi and Rehema Mgoda for their support as research assistance. Lastly but not least, we thank you all people who have participated and contributed to this study.

## Authors’ Information

1. Henry A. Mollel (Corresponding Author) Affiliation: Department of Health Information Systems, Mzumbe University, Morogoro, Tanzania.
2. Lawrencia D. Mushi Affiliation: Department of Health Information Systems, Mzumbe University, Morogoro, Tanzania.
3. Richard V. Nkwera Affiliation: Centre of Excellence in Health Monitoring and Evaluation, Mzumbe University, Morogoro, Tanzania.

